# Phenotypes of cough in children: a latent class analysis

**DOI:** 10.1101/2023.03.09.23287047

**Authors:** Maria Christina Mallet, Eva SL Pedersen, Ronny Makhoul, Sylvain Blanchon, Karin Hoyler, Anja Jochmann, Philipp Latzin, Alexander Moeller, Nicolas Regamey, SPAC Study Team, Myrofora Goutaki, Ben D Spycher, Claudia E Kuehni

**Affiliations:** Institute of Social and Preventive Medicine, University of Bern, Bern, Switzerland; Graduate School for Health Sciences, University of Bern, Bern, Switzerland; Dept Woman-Mother-Child, Service of Pediatrics, Pediatric Pulmonology and Cystic Fibrosis Unit, Lausanne University Hospital and University of Lausanne, Lausanne, Switzerland; Kinderpneumologie Horgen, Private Practice for Pediatric Pneumology, Horgen, Switzerland; Department of Paediatric Pulmonology, University Children’s Hospital Basel, Switzerland; Division of Paediatric Respiratory Medicine and Allergology, Department of Paediatrics, Inselspital, Bern University Hospital, University of Bern, Bern, Switzerland; Department of Respiratory Medicine, University Children’s Hospital Zurich and Children’s Research Centre, University of Zurich, Zurich, Switzerland; Division of Paediatric Pulmonology, Children’s Hospital, Cantonal Hospital Lucerne, Switzerland

**Keywords:** allergy, childhood, clinical phenotypes, cough, latent class analysis, FeNO, respiratory symptoms, spirometry, unsupervised learning

## Abstract

**Background:** Distinguishing phenotypes among children with cough helps understand underlying causes. Using a statistical data-driven approach, we identified cough phenotypes and we aimed to validate them based on measurable traits, physician diagnoses, and prognosis.

**Methods:** We used data from 531 children aged 5–16 years from the Swiss Paediatric Airway Cohort—a multicentre clinical cohort of children seen in outpatient clinics since 2017. We included children with any parent-reported cough (i.e. cough without a cold, cough at night, cough more than others, or cough longer than 4 weeks) without current wheeze. We applied latent class analysis to identify phenotypes using 9 symptoms and characteristics and selected the best model using the Akaike Information Criterion. We assigned children to the most likely phenotype and compared the resulting groups with regards to parental history, comorbidities, measurable traits, physician diagnoses, and prognosis after 1 year.

**Results:** Our analysis distinguished 4 cough phenotypes: 1. unspecific dry cough (25%); 2. non-allergic infectious and night cough with snoring and otitis (4%); 3. allergic dry night cough with snoring (9%); and 4. allergic cough (61%). Children with the allergic phenotype often had family or personal history of atopy and were diagnosed with asthma. Fractional exhaled nitric oxide was highest for the allergic phenotype [median 17.9 parts per billion (ppb)] and lowest for the non-allergic infectious phenotype (median 7.0 ppb). Positive allergy test results differed across phenotypes (p<0.001) and were most common among the allergic (70%) and least common among the unspecific dry cough (31%) phenotypes. Subsequent wheeze was thrice as high among the allergic than the unspecific dry cough phenotype.

**Conclusion:** We distinguished 4 clinically-relevant cough phenotypes; they differed by measurable traits, physician diagnoses, and prognosis. Although we excluded children with current wheeze, most children belonged to allergy-related phenotypes and possibly need allergy and asthma work-ups.

**Three key messages:**

1. Latent class analysis identified 4 cough phenotypes distinguished by measurable traits, diagnoses, and prognosis.
2. Most children belonged to allergy-related phenotypes and possibly need allergy and asthma work-ups.
3. Symptom-based cough phenotypes correlate with measurable traits and potentially apply to all healthcare settings.

## 1. INTRODUCTION

Recurrent cough is a frequent symptom in paediatric care and the underlying aetiologies are heterogeneous.^[1-3]^ Most common causes of cough include respiratory tract infections and asthma, yet many children cough in the absence of wheeze, colds, or obvious underlying disease, which presents diagnostic challenges for physicians.^[1, 3-5]^ Many causes have been described for the latter group of children, ranging from upper airway cough syndrome caused by allergic and non-allergic rhinitis, asthma–like conditions and gastroesophageal reflux to psychogenic and habit/tic cough disorders.^[1, 6-10]^ Clinical guidelines for differential diagnosis of cough among children are used to classify cough into aetiologies based on cough duration, dry versus wet cough, triggers, associated symptoms and diseases, and measurable physiological traits, such as lung function and allergy tests. These guidelines are usually based on expert opinions and involve some degree of overlap between diagnoses.^[11- 13]^ Unsupervised learning methods, such as latent class analysis (LCA), are increasingly being used as a more objective, data-driven approach to distinguish phenotypes of disease. Such methods are not limited by predefined categories and account for multiple disease dimensions.^[14, 15]^ Distinguishing phenotypes may lead to new hypotheses concerning underlying aetiologies and mechanisms, ease diagnostic work-ups, guide targeted management, and predict outcomes, such as respiratory symptoms.^[15-17]^

Although LCA is widely used to classify children with wheeze, asthma, or bronchiolitis, ^[18-25]^ few have applied LCA to children presenting with recurrent cough.^[15, 26-28]^ Present studies were population-based, thus mainly included children with rather mild disease, many of whom had not seen a paediatric respiratory specialist for cough. In addition, most studies were either focused on asthma and wheeze with little emphasis on cough—thus limiting identification of cough phenotypes^[15, 26, 27]^—or included measurable physiological traits for phenotype definition, which limits use across healthcare settings with restricted or no access to diagnostic tests.^[15, 26]^ We aimed to identify phenotypes of cough in children consulting physicians, using information that is readily available in all healthcare settings, including primary care. Using data from a clinical population of children presenting with cough without current wheeze, we applied LCA to a set of cough-related symptoms and characteristics to identify and describe cough phenotypes. We further aimed to validate the identified phenotypes by assessing their associations with clinically-relevant features, including parent-reported information, paediatric pulmonologist diagnoses, measurable physiological traits, and prognosis of respiratory symptoms 1 year later.

## 2. METHODS

### 2.1 Study design and population

The Swiss Paediatric Airway Cohort (SPAC) is a prospective clinical study integrated into the routine care of 8 paediatric respiratory outpatient clinics and 2 paediatric practices across Switzerland. SPAC design and methods are described in detail elsewhere.^[29]^ SPAC enrols children aged 0–16 years referred by general practitioners or paediatricians for the evaluation of common respiratory problems, such as cough, wheeze, and exercise-induced symptoms. At time of referral, parents or guardians complete a baseline questionnaire about respiratory symptoms, socio-demographic information, environmental exposures, personal and family history, and comorbidities. One year after completing the baseline questionnaire and yearly thereafter, parents receive a follow-up questionnaire asking about symptoms and treatments during the past 12 months. From hospital records, we retrieve information about results of diagnostic tests and paediatric pulmonologist diagnoses. The Bern Cantonal Ethics Committee (Kantonale Ethikkomission Bern 2016-02176) approved the study; parents and patients older than 14 years provided their written consent.

### 2.2 Inclusion criteria

We only included children aged 5–16 years at the time of visit because physiological traits, such as lung function and allergies, are routinely measured for this age group. We included all children with completed baseline parental questionnaires by December 31, 2020 who indicated suffering from any recurrent cough. We used 4 validated and commonly used questions about cough: “Does your child have a cough even without having a cold?” (cough without a cold);^[30]^ “In the last 12 months has your child had a dry cough at night, apart from a cough associated with a cold or a chest infection?” (dry night cough);^[31]^ “Do you think your child coughs more than other children?” (cough more than others);^[32]^ and “Has your child had a cough in the past 12 months that lasted more than 4 weeks in a row?” (chronic cough). We excluded children who reported wheeze in the past 12 months (current wheeze), but not those who reported wheeze earlier in life (> 12 months ago) or with physician-diagnosed asthma earlier in life.

### 2.3 Definition of indicator variables

As indicator variables for the latent class model, we included the following 9 variables, all reported during the last 12 months from the baseline questionnaire: frequency of colds (≥ 7/year), pneumonia, otitis media, regular snoring, rhino-conjunctivitis, dry night cough, type of cough (dry versus wet), duration of cough (> 2 months), and number of allergic cough triggers, including pets, pollen, and house dust, ranging from 0 to 3 (Table S1). We selected these cough-related symptoms and characteristics based on a consensus among authors as they were considered to be potential indicators of underlying aetiologies that can be easily asked in all clinical settings, including primary care.

### 2.4 Definition of variables to characterise and validate identified phenotypes

We characterised and validated the identified phenotypes by comparing how these differed according to a) parent-reported information, b) paediatric pulmonologist diagnoses, c) results of measurable traits, and d) prognosis of respiratory symptoms 1 year later. First, we compared the phenotypes with respect to exposure, family history, and comorbidity information reported by parents in the baseline questionnaire. Second, we compared the phenotypes with diagnoses given by paediatric pulmonologists who were unaware of the LCA results. For simplicity, we grouped diagnoses into broad categories— asthma, chronic cough, allergies, and others. A child could have several diagnoses. Appendix S1 provides details of parent-reported information and physician diagnoses. Third, we validated the identified phenotypes against measurable traits. We used body mass index (BMI), spirometry, bronchodilator response (BDR), fractional exhaled nitric oxide (FeNO), and allergy tests [skin prick test (SPT) and allergen specific IgE (sIgE)]. We converted BMI into z-scores using reference values from the World Health Organisation.^[33]^ For spirometry, we transformed forced vital capacity (FVC), forced expiratory volume in the first second (FEV1), FEV1/FVC, and the forced expiratory flow between the 25% and 75% of the FVC (FEF 25–75) into z-scores using the Global Lung Initiative reference values.^[34]^ We defined a positive BDR test as an increase in FEV1 ≥ 12% (from baseline FEV1) after the use of a short acting beta-agonist. FeNO measurements differed across clinics, with cut-offs varying from 10 parts per billion (ppb) when using an offline method to 20–25 ppb for online methods.^[35]^ Thus, we first distinguished low versus high FeNO based on specified cut- offs for each centre. We also assessed FeNO (measured online) as a continuous variable and expressed in ppb. A positive allergy test was defined as either a positive SPT (mean wheal size of ≥ 3mm) or a positive sIgE with values ≥ 0.35 kU/L. Since SPAC is an observational study, physiological measurements are performed based on local protocols and indications and not for all children. However, availability of measurements did not differ across phenotypes (Table S2).

Fourth, we compared prognosis 1 year later between phenotypes by looking at reported cough without a cold, dry night cough, cough more than others, cough more than 4 weeks, and wheeze during the past 12 months. Parents of 360 (68%) children completed the first year follow-up questionnaire and response rates did not differ by phenotype (Table S2).

### 2.5 Statistical analysis

To identify different phenotypes of cough, we fitted latent class models to the dataset including the selected cough-related variables. To select number of latent classes (interpreted here as phenotypes), we repeatedly fitted the model with the number of classes increasing stepwise from 2 to 10 and used Bayesian Information Criterion (BIC) and Akaike Information Criterion (AIC) to select the optimal model.^[36]^ These criteria favour models that are parsimonious, i.e. have few model parameters (in our context this means fewer classes), while still achieving good model fit. For the final selection, we also took into account interpretability and clinical relevance. We fitted models using the generalised structural equation modelling function (gsem) in STATA. The dichotomised symptom variables were assumed to be binomially distributed within classes and the number of allergic triggers as Poisson distributed.

For validation of phenotypes, we first assigned each individual to the phenotypes for which it had the highest posterior membership probability.^[14]^ We then compared the resulting groups with respect to parent-reported characteristics, paediatric pulmonologists diagnoses, measured traits and prognosis of respiratory symptoms after 1 year. We tested differences between phenotypes using chi-squared and Fisher exact tests for categorical variables. For non-normally distributed continuous variables we used Kruskal –Wallis tests.

We recoded missing values for questions about symptoms as “no” since we assumed absent or mild symptoms when parents have not answered “yes” (Table S1). We performed all analyses in STATA (Version 15.1, StataCorp, TX, USA).

## 3. RESULTS

### 3.1 Characteristics of study population

Our analysis included 531 children with median age 11 years (interquartile range 8–13) (Figure S1, Table 1). Parents of 446 (84%) children reported cough without a cold, 289 (54%) cough more than others, 276 (52%) dry night cough, and 215 (40%) cough more than 4 weeks.

**Table 1:** Characteristics of study population included in the latent class analysis at baseline (N=531)

|  | N (%) |
| --- | --- |
| **Sociodemographic and environmental exposure** |  |
| Male sex | 314 (59) |
| Age, years (median, IQR) | 11 (8–13) |
| Overweight or obesity based on BMI z-scores (N=521) | 125 (24) |
| Swiss nationality | 436 (82) |
| Parental highest education level |  |
| Maternal tertiary (N=511) | 149 (29) |
| Paternal tertiary (N=504) | 203 (40) |
| Exposure to tobacco smoke (N=498) | 142 (29) |
| **Parental history** |  |
| Asthma or asthma inhaler use | 191 (36) |
| Hay fever | 247 (47) |
| Chronic cough | 73 (14) |
| **Comorbidities ever in life** |  |
| Previous wheeze | 223 (42) |
| Physician-diagnosed asthma | 245 (46) |
| Hay fever | 222 (42) |
| Physician-diagnosed eczema | 112 (21) |
| **Cough symptoms and characteristics during the past 12 months** |  |
| Dry night cough | 276 (52) |
| Cough without a cold | 446 (84) |
| Cough more than others | 289 (54) |
| Cough duration > 4 weeks | 215 (40) |
| Cough duration > 2 months | 79 (15) |
| Cough type |  |
| Mostly dry | 253 (48) |
| Mostly wet | 53 (10) |
| Both dry and wet | 221 (42) |
| Cough triggered by any aero-allergens (pollen, house dust, animals) | 318 (60) |
| **Other symptoms in the past 12 months** |  |
| Number of colds ≥ 7/year | 61 (11) |
| Otitis media | 47 (9) |
| Pneumonia | 31(6) |
| Rhino-conjunctivitis | 195 (37) |
| Snoring (without cold/almost always) | 186 (35) |

IQR: Interquartile range, BMI: Body Mass Index, we calculated BMI z-scores using reference values from the World Health Organisation; Parental history, comorbidities ever, cough and other symptoms are parent-reported.

### 3.2 Description of phenotypes

Results of BIC and AIC were discordant: the lowest BIC indicated a model with 2 phenotypes and the lowest AIC a model with 4 phenotypes (Table S3). Since identifying cough phenotypes using LCA is exploratory, we present the model with 4 phenotypes which we think is more clinically-relevant, assuming there are more than 2 underlying causes for recurrent cough among children^[13]^. We summarise class size, main characteristics, and summary labels of the 4 phenotypes in the textbox, Figure 1, and Table S4.

**Figure 1:**
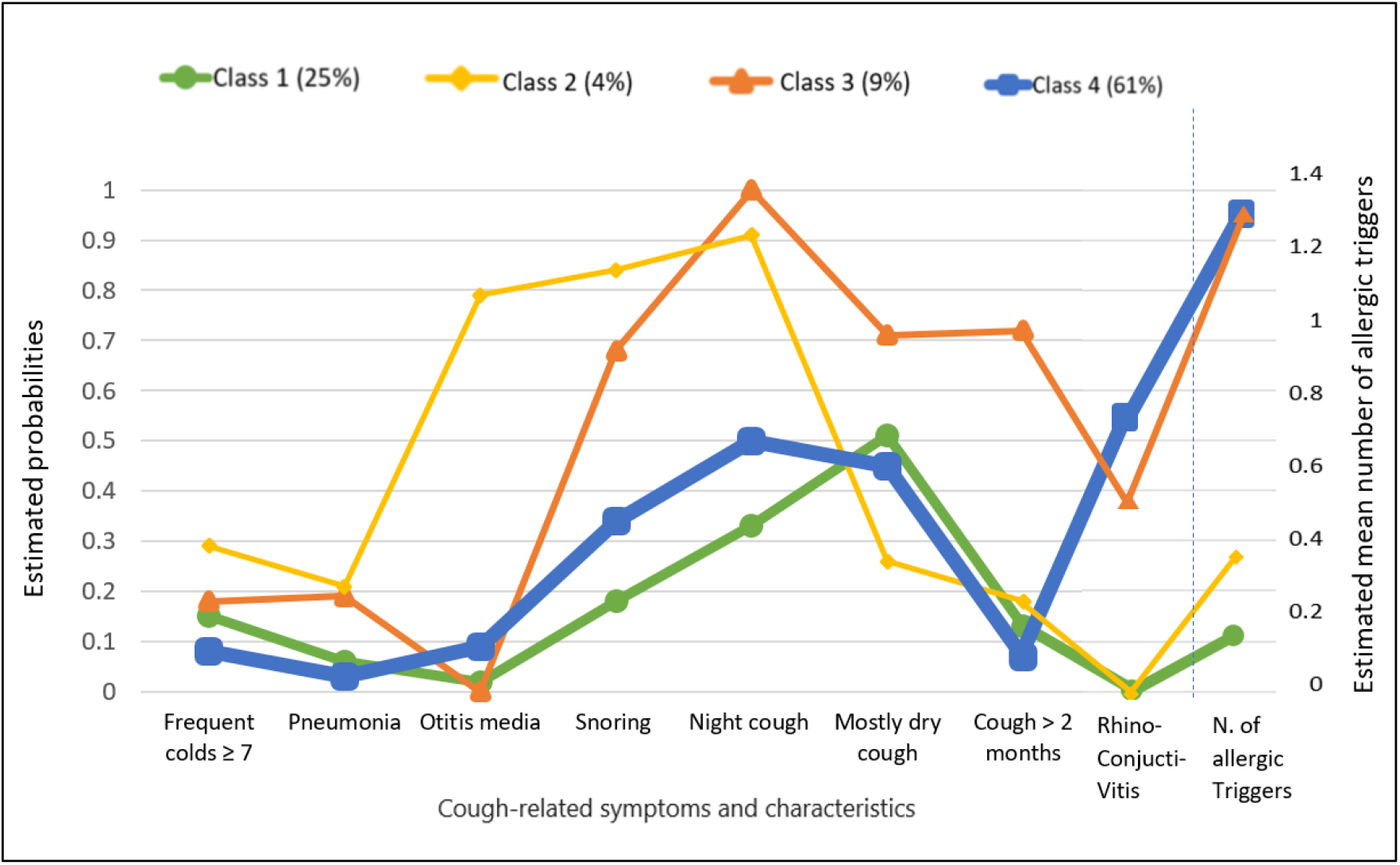
Estimated class sizes (in brackets)* and estimated probabilities of cough-related symptoms and characteristics of the four-class model identified by latent class analysis based on the lowest Akaike Information Criterion (N=531) *The width of the lines represents the size of the classes, i.e. the proportion of children in the different classes. The estimated probabilities which can take values from 0 to 1 and represent the probability of answering ‘yes’ for the dichotomous indicators variables (frequent colds ≥ 7, pneumonia, otitis media, night cough, rhino-conjunctivitis, snoring, mostly dry cough, cough > 2 months) for each of the 4 classes, while the estimated mean which can take values from 0 to 3 represents the average number of allergic cough triggers for each of the 4 classes.

**4 cough phenotypes identified by latent class analysis** (Details in Table S4)

*Phenotype 1: Unspecific dry cough (25%)*

Children belonging to this phenotype reported few respiratory infections or allergic symptoms. Cough was described as mostly dry for half of the children without any other distinct characteristics.

*Phenotype 2: Non-allergic infectious and night cough with snoring and otitis (4%)*

Frequent colds and pneumonia were more frequently reported than in other phenotypes. Children in this phenotype typically reported otitis media, night cough, and snoring. None had rhino-conjunctivitis, hardly any reported allergic triggers for cough, and cough was rarely reported as dry.

*Phenotype 3: Allergic dry night cough with snoring (9%)*

Night cough, snoring, dry cough, and cough lasting more than two consecutive months were typical for this phenotype. Rhino-conjunctivitis and allergic cough triggers were much more commonly reported compared with phenotypes 1 and 2.

*Phenotype 4: Allergic cough (61%)*

Nearly two-thirds of children were in this phenotype with highest probabilities of rhino- conjunctivitis and highest mean number of reported allergic triggers for cough. Very few colds, pneumonia, or otitis media were reported. The probability of reporting night cough and snoring was half of phenotype 3.

For completeness, we also present the model with 2 phenotypes in Table S5; it identified a larger phenotype (73% of children) with allergic features (rhino-conjunctivitis, allergic triggers), as the four-class model, and a smaller, less specific phenotype.

### 3.3 Validation of identified phenotypes

Parent-reported characteristics, paediatric pulmonologist diagnoses, measurable traits, and respiratory prognosis varied between the different phenotypes. Children with phenotype 2 were more often male (72%) and younger (median age: 7 years) compared with other phenotypes (Table 2). Family history of asthma and hay fever and atopy-related comorbidities, such as previous wheeze, physician-diagnosed asthma, hay fever, and eczema, were reported for all phenotypes but were most common among children with phenotype 4 (allergic cough).

**Table 2:**
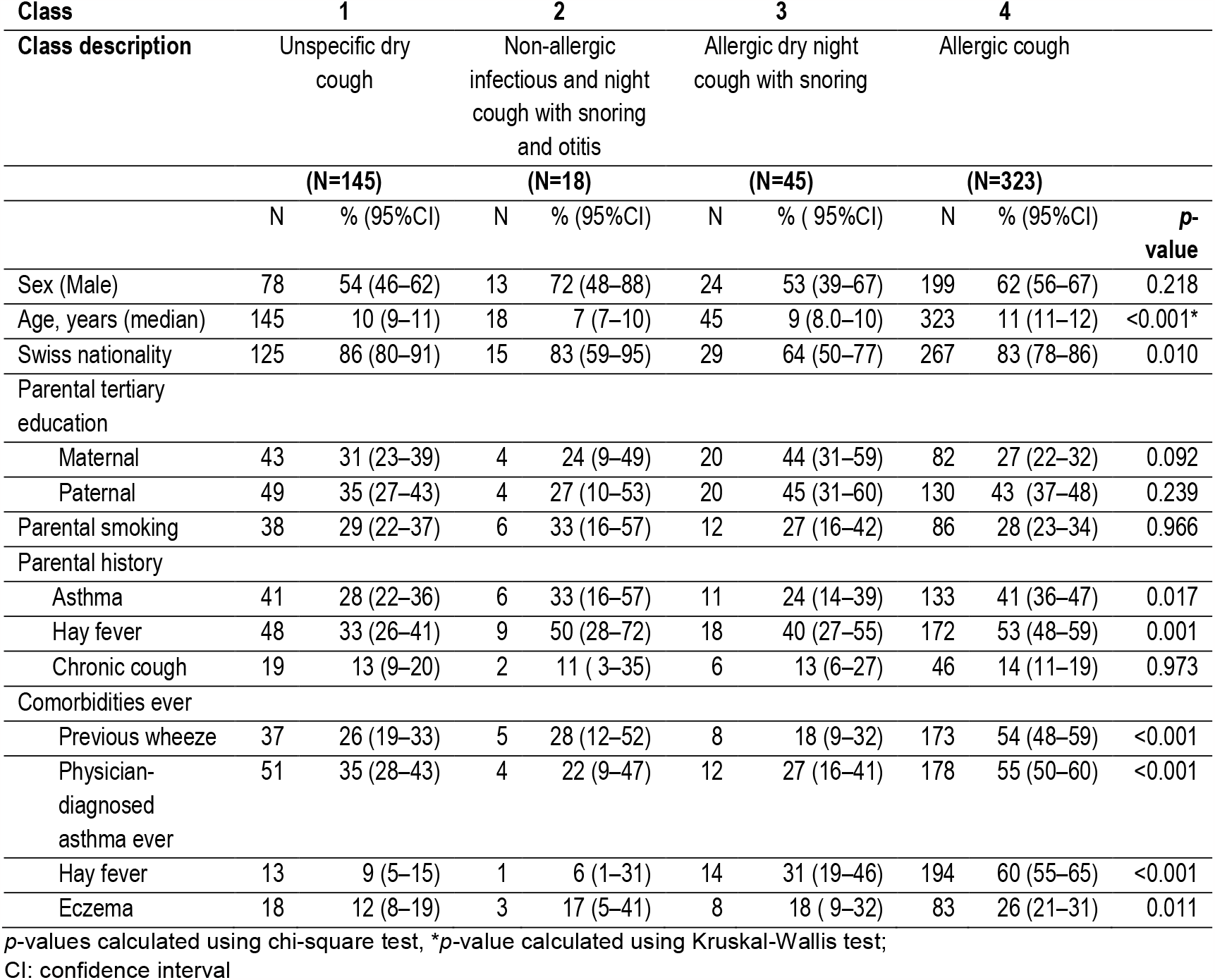
Socio-demographic characteristics, parental smoking, family history, and comorbidities for the 4 cough phenotypes (N=531)

| Class | 1 |  | 2 |  | 3 |  | 4 |  |  |
| --- | --- | --- | --- | --- | --- | --- | --- | --- | --- |
| Class description | Unspecific dry cough |  | Non-allergic infectious and night cough with snoring and otitis |  | Allergic dry night cough with snoring |  | Allergic cough |  |  |
|  | (N=145) |  | (N=18) |  | (N=45) |  | (N=323) |  |  |
|  | N | % (95%CI) | N | % (95%CI) | N | % (95%CI) | N | % (95%CI) | p-value |
| Sex (Male) | 78 | 54 (46–62) | 13 | 72 (48–88) | 24 | 53 (39–67) | 199 | 62 (56–67) | 0.218 |
| Age, years (median) | 145 | 10 (9–11) | 18 | 7 (7–10) | 45 | 9 (8.0–10) | 323 | 11 (11–12) | <0.001* |
| Swiss nationality | 125 | 86 (80–91) | 15 | 83 (59–95) | 29 | 64 (50–77) | 267 | 83 (78–86) | 0.010 |
| Parental tertiary education |  |  |  |  |  |  |  |  |  |
| Maternal | 43 | 31 (23–39) | 4 | 24 (9–49) | 20 | 44 (31–59) | 82 | 27 (22–32) | 0.092 |
| Paternal | 49 | 35 (27–43) | 4 | 27 (10–53) | 20 | 45 (31–60) | 130 | 43 (37–48) | 0.239 |
| Parental smoking | 38 | 29 (22–37) | 6 | 33 (16–57) | 12 | 27 (16–42) | 86 | 28 (23–34) | 0.966 |
| Parental history |  |  |  |  |  |  |  |  |  |
| Asthma | 41 | 28 (22–36) | 6 | 33 (16–57) | 11 | 24 (14–39) | 133 | 41 (36–47) | 0.017 |
| Hay fever | 48 | 33 (26–41) | 9 | 50 (28–72) | 18 | 40 (27–55) | 172 | 53 (48–59) | 0.001 |
| Chronic cough | 19 | 13 (9–20) | 2 | 11 (3–35) | 6 | 13 (6–27) | 46 | 14 (11–19) | 0.973 |
| Comorbidities ever |  |  |  |  |  |  |  |  |  |
| Previous wheeze | 37 | 26 (19–33) | 5 | 28 (12–52) | 8 | 18 (9–32) | 173 | 54 (48–59) | <0.001 |
| Physician-diagnosed asthma ever | 51 | 35 (28–43) | 4 | 22 (9–47) | 12 | 27 (16–41) | 178 | 55 (50–60) | <0.001 |
| Hay fever | 13 | 9 (5–15) | 1 | 6 (1–31) | 14 | 31 (19–46) | 194 | 60 (55–65) | <0.001 |
| Eczema | 18 | 12 (8–19) | 3 | 17 (5–41) | 8 | 18 (9–32) | 83 | 26 (21–31) | 0.011 |

p-values calculated using chi-square test, \*p-value calculated using Kruskal-Wallis test;
CI: confidence interval

Paediatric pulmonologist diagnoses differed across the 4 phenotypes (Table 3). Asthma diagnosis was most frequent in phenotype 4 (48%), followed by phenotype 1 (26%), phenotype 3 (20%), and phenotype 2 (6%). Phenotype 3 contained the highest proportion of children with a specific cough diagnosis (33%) by a paediatric pulmonologist, whilst in phenotype 4 this number was smallest (8%). Children with phenotype 4 more often received a diagnosis of rhinitis/rhino-conjunctivitis and allergic sensitisation/atopy.

**Table 3:** Diagnoses given by physicians to children in the 4 cough phenotypes (N=531)

| **Class** |  | **1** | **2** | **3** | **4** |  |
| --- | --- | --- | --- | --- | --- | --- |
| **Class description** | Total | Unspecific dry cough | Non-allergic infectious and night cough with snoring and otitis | Allergic dry night cough with snoring | Allergic cough |  |
|  | (N=531) | (N=145) | (N=18) | (N=45) | (N=323) |  |
|  | **N (%)** | **N (%)** | **N (%)** | **N (%)** | **N (%)** | **p-value** |
| Confirmed asthma<sup>1</sup> | 204 (38) | 38 (26) | 1 (6) | 9 (20) | 156 (48) | <0.001 |
| Any confirmed or suspected asthma/asthma-like diagnosis<sup>2</sup> | 370 (70) | 80 (55) | 8 (44) | 22 (49) | 260 (81) | <0.001 |
| Any confirmed chronic cough diagnosis<sup>3</sup> | 73 (14) | 29 (20) | 4 (22) | 15 (33) | 25 (8) | <0.001 |
| Any confirmed or suspected chronic cough diagnosis<sup>4</sup> | 103 (19) | 41 (28) | 7 (39) | 20 (44) | 35 (11) | <0.001 |
| Any confirmed exercise-related problem<sup>5</sup> | 22 (4) | 10 (7) | 2 (11) | 1 (2) | 9 (3) | 0.059* |
| Any confirmed or suspected exercise-related problem<sup>6</sup> | 46 (9) | 20 (14) | 2 (11) | 2 (4) | 22 (7) | 0.063* |
| Any confirmed or suspected chronic snoring problem<sup>7</sup> | 24 (5) | 7 (5) | 2 (11) | 1 (2) | 14(4) | 0.453* |
| Confirmed rhinitis/rhino-conjunctivitis | 213 (40) | 11 (8) | 3 (17) | 15 (33) | 184 (57) | <0.001 |
| Confirmed allergic sensitisation/atopy | 351 (66) | 48 (33) | 8 (44) | 28 (62) | 267 (83) | <0.001 |

Note: A child can have multiple diagnosis. *p*-values calculated using chi-square test \**p*-value calculated using Fisher's exact test
<sup>1</sup> *confirmed* diagnosis of asthma (chronic asthma)
<sup>2</sup> *confirmed or suspected* diagnosis of asthma (episodic asthma, chronic asthma, exercise-induced asthma) and asthma-like diagnosis (wheeze, recurrent bronchitis, bronchial hyperreactivity, exercise-induced wheeze)
<sup>3</sup> *any confirmed* diagnosis of chronic cough, including recurrent cough of unknown aetiology, increased vulnerability to viral infections, post infectious cough, psychogenic cough, chronic obstructive rhinopathy with posterior rhinorrhoea /postnasal drip, cough hypersensitivity syndrome, protracted bacterial bronchitis, gastroesophageal reflux disease
<sup>4</sup> *any confirmed or suspected* diagnosis of chronic cough, including recurrent cough of unknown aetiology, increased vulnerability to viral infections, post infectious cough, psychogenic cough, chronic obstructive rhinopathy with posterior rhinorrhoea /postnasal drip, cough hypersensitivity syndrome, protracted bacterial bronchitis, gastroesophageal reflux disease
<sup>5</sup> *any confirmed* exercise-related problems, including dysfunctional breathing, hyperventilation syndrome, stridor, sighing, vocal cord dysfunction, induced laryngeal obstruction, deconditioning
<sup>6</sup> *any confirmed or suspected* exercise-related problem: dysfunctional breathing, hyperventilation syndrome, stridor, sighing, vocal cord dysfunction, induced laryngeal obstruction, deconditioning
<sup>7</sup> *any confirmed or suspected* chronic snoring problem, including adenoid hypertrophy, tonsil hypertrophy, obstructive sleep apnea, unspecific snoring

Regarding measurable traits, BMI did not differ across the 4 phenotypes (*p*=0.161) (Table 4). Median z-scores of spirometry parameters (FCV, FEV1/FVC, and FEF25-75) were lowest for phenotype 2 and highest for phenotype 3 (Table 4, Figures 2A and S2). Fewer than half the children had a BDR test done and among those, the proportion with a positive BDR test did not differ across phenotypes (p=0.406). FeNO values differed across groups (p<0.001) being highest among children with phenotype 4 [17.9 ppb, 95% confidence interval (CI) 15.6–19.5] and lowest for children with phenotype 2 (7.0 ppb, 95% CI 5.0–17.7) (Table 4, Figure 2B). Similarly, positive allergy tests differed across phenotypes (p< 0.001) and were more common among children with phenotype 4 (70%, 95% CI 64–76), and relatively rare for children with phenotype 1 (31%, 95% CI 23–41).

**Table 4:**
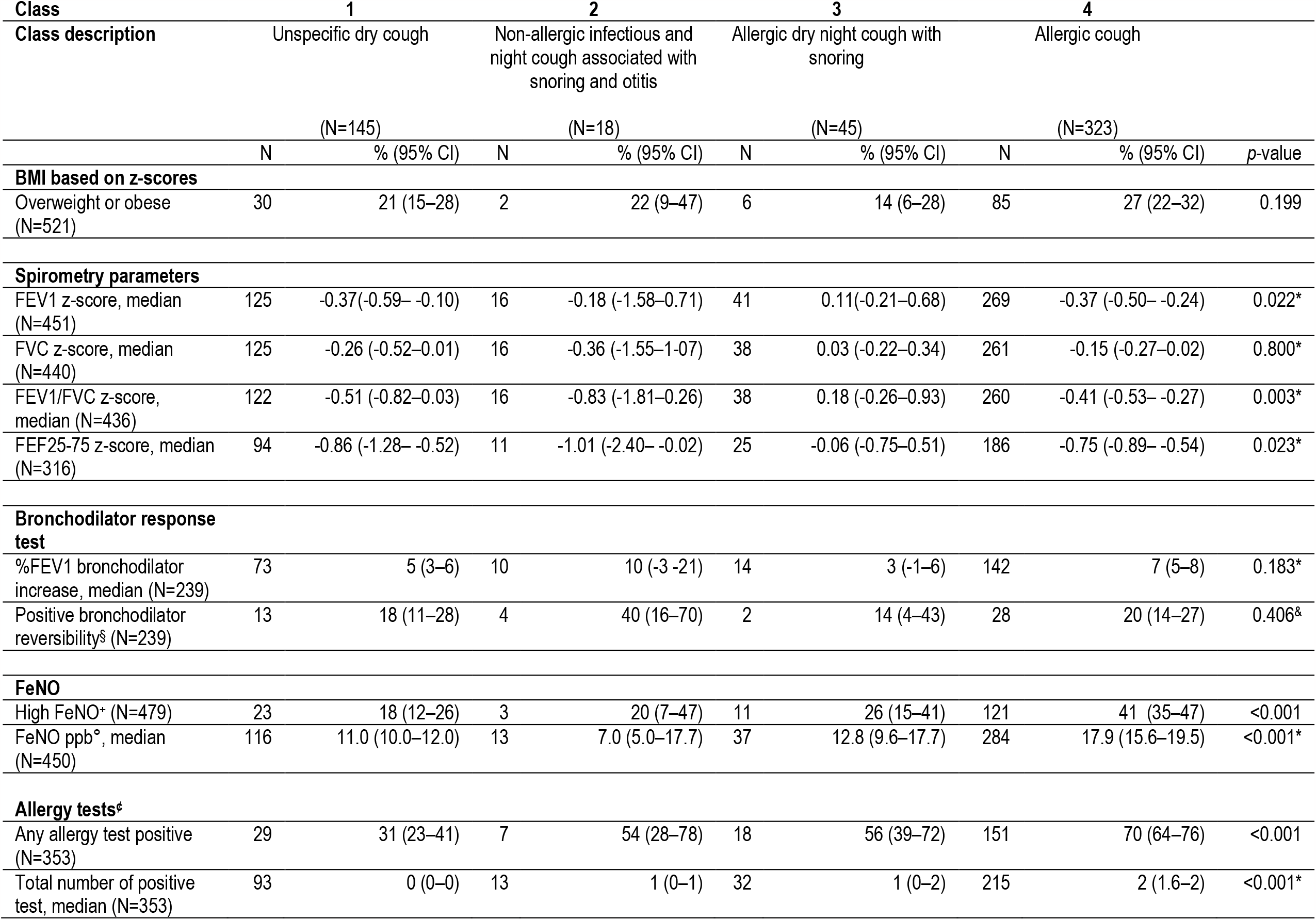
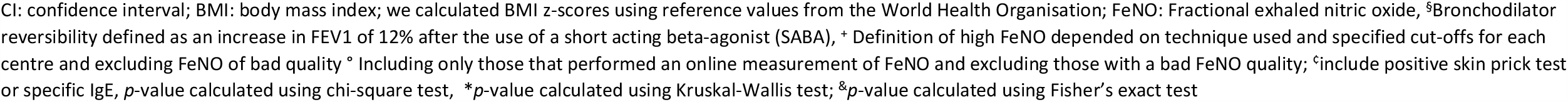
Comparison of objective features and physiological measurements based on the 4 identified cough phenotypes

| Class | 1 |  | 2 |  | 3 |  | 4 |  |  |
| --- | --- | --- | --- | --- | --- | --- | --- | --- | --- |
| Class description | Unspecific dry cough |  | Non-allergic infectious and night cough associated with snoring and otitis |  | Allergic dry night cough with snoring |  | Allergic cough |  |  |
|  | (N=145) |  | (N=18) |  | (N=45) |  | (N=323) |  |  |
|  | N | % (95% CI) | N | % (95% CI) | N | % (95% CI) | N | % (95% CI) | p-value |
| **BMI based on z-scores** |  |  |  |  |  |  |  |  |  |
| Overweight or obese (N=521) | 30 | 21 (15–28) | 2 | 22 (9–47) | 6 | 14 (6–28) | 85 | 27 (22–32) | 0.199 |
| **Spirometry parameters** |  |  |  |  |  |  |  |  |  |
| FEV1 z-score, median (N=451) | 125 | -0.37 (-0.59– -0.10) | 16 | -0.18 (-1.58–0.71) | 41 | 0.11 (-0.21–0.68) | 269 | -0.37 (-0.50– -0.24) | 0.022* |
| FVC z-score, median (N=440) | 125 | -0.26 (-0.52–0.01) | 16 | -0.36 (-1.55–1.07) | 38 | 0.03 (-0.22–0.34) | 261 | -0.15 (-0.27–0.02) | 0.800* |
| FEV1/FVC z-score, median (N=436) | 122 | -0.51 (-0.82–0.03) | 16 | -0.83 (-1.81–0.26) | 38 | 0.18 (-0.26–0.93) | 260 | -0.41 (-0.53– -0.27) | 0.003* |
| FEF25-75 z-score, median (N=316) | 94 | -0.86 (-1.28– -0.52) | 11 | -1.01 (-2.40– -0.02) | 25 | -0.06 (-0.75–0.51) | 186 | -0.75 (-0.89– -0.54) | 0.023* |
| **Bronchodilator response test** |  |  |  |  |  |  |  |  |  |
| %FEV1 bronchodilator increase, median (N=239) | 73 | 5 (3–6) | 10 | 10 (-3–21) | 14 | 3 (-1–6) | 142 | 7 (5–8) | 0.183* |
| Positive bronchodilator reversibility<sup>§</sup> (N=239) | 13 | 18 (11–28) | 4 | 40 (16–70) | 2 | 14 (4–43) | 28 | 20 (14–27) | 0.406<sup>§</sup> |
| **FeNO** |  |  |  |  |  |  |  |  |  |
| High FeNO* (N=479) | 23 | 18 (12–26) | 3 | 20 (7–47) | 11 | 26 (15–41) | 121 | 41 (35–47) | <0.001 |
| FeNO ppb°, median (N=450) | 116 | 11.0 (10.0–12.0) | 13 | 7.0 (5.0–17.7) | 37 | 12.8 (9.6–17.7) | 284 | 17.9 (15.6–19.5) | <0.001* |
| **Allergy tests<sup>‡</sup>** |  |  |  |  |  |  |  |  |  |
| Any allergy test positive (N=353) | 29 | 31 (23–41) | 7 | 54 (28–78) | 18 | 56 (39–72) | 151 | 70 (64–76) | <0.001 |
| Total number of positive test, median (N=353) | 93 | 0 (0–0) | 13 | 1 (0–1) | 32 | 1 (0–2) | 215 | 2 (1.6–2) | <0.001* |

CI: confidence interval; BMI: body mass index; we calculated BMI z-scores using reference values from the World Health Organisation; FeNO: Fractional exhaled nitric oxide, <sup>§</sup>Bronchodilator reversibility defined as an increase in FEV1 of 12% after the use of a short acting beta-agonist (SABA), <sup>+</sup> Definition of high FeNO depended on technique used and specified cut-offs for each centre and excluding FeNO of bad quality <sup>°</sup> Including only those that performed an online measurement of FeNO and excluding those with a bad FeNO quality; <sup>°</sup>include positive skin prick test or specific IgE, *p*-value calculated using chi-square test, <sup>\*</sup>*p*-value calculated using Kruskal-Wallis test; <sup>&</sup>*p*-value calculated using Fisher's exact test

**Figure 2.**
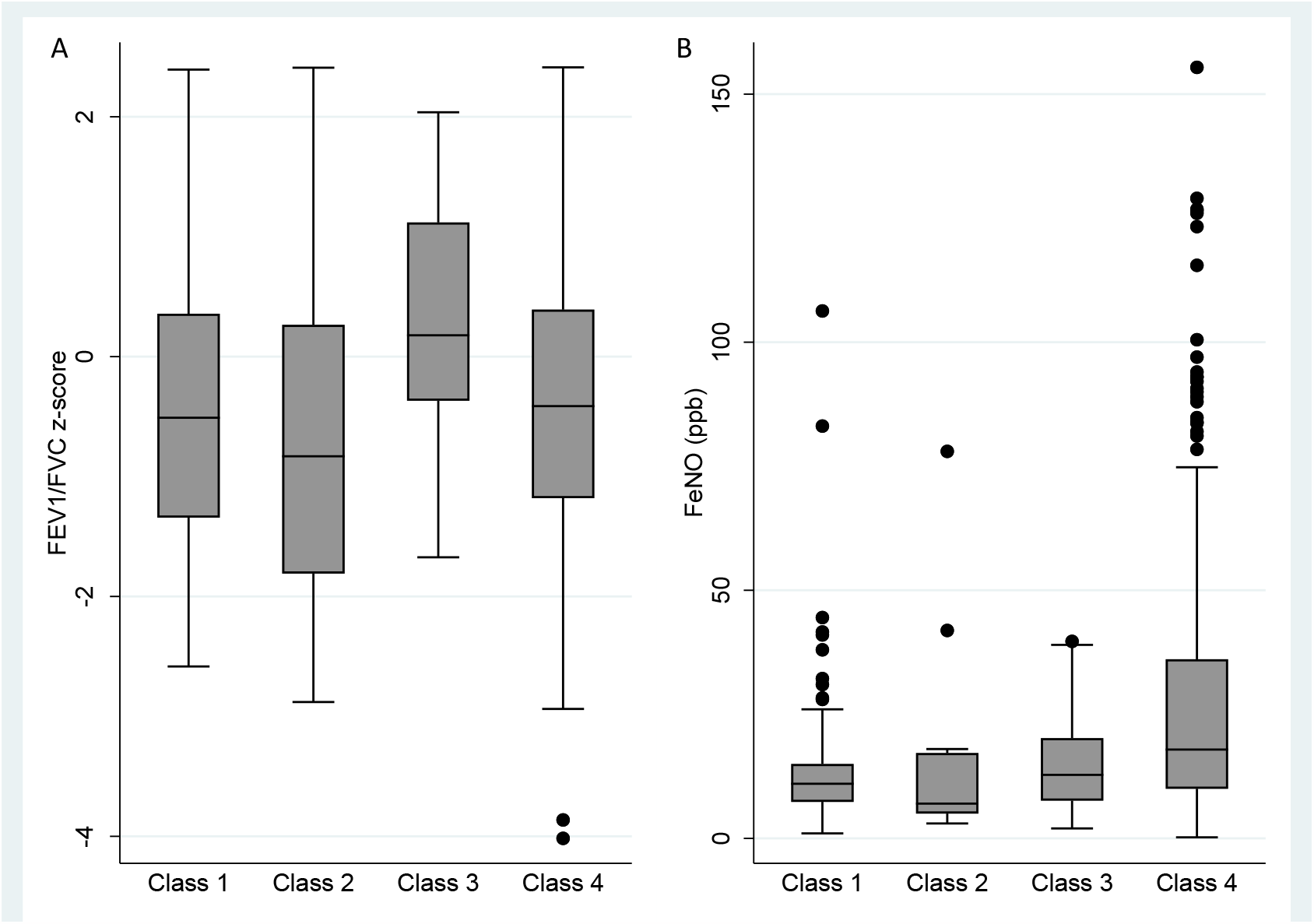
**(A):** Distribution of spirometry parameters FEV1/FVC z-score (N=436) and **(B)** Distribution of FeNO values based on the 4 identified cough phenotypes (N=450) FEV1: Forced Expiratory Volume in one second; FVC: Forced Vital Capacity; FeNO: Fractional Exhaled Nitric Oxide; ppb: parts per billion; include FeNO values measured using the online method **Class 1**: Unspecific dry cough **Class 2**: Non-allergic infectious and night cough with snoring and otitis **Class 2:** Allergic dry night cough with snoring **Class 4**: Allergic cough

Regarding prognosis after 1 year, overall, cough was less often reported at follow-up (Figure 3) compared with baseline (Table 1). Children with phenotype 3 more often reported cough without a cold (66%), cough more than others (52%), and cough more than 4 weeks (24%) compared with children in other phenotypes (Figure 3). Night cough was more prevalent for children with phenotype 2 (57%) followed by children with phenotype 3 (41%). Wheeze was reported among 78 (22%) children and the proportion differed across the 4 phenotypes (p=0.005). Children with phenotype 4 wheezed almost 3 times more after 1 year compared with children in phenotype 1.

**Figure 3:**
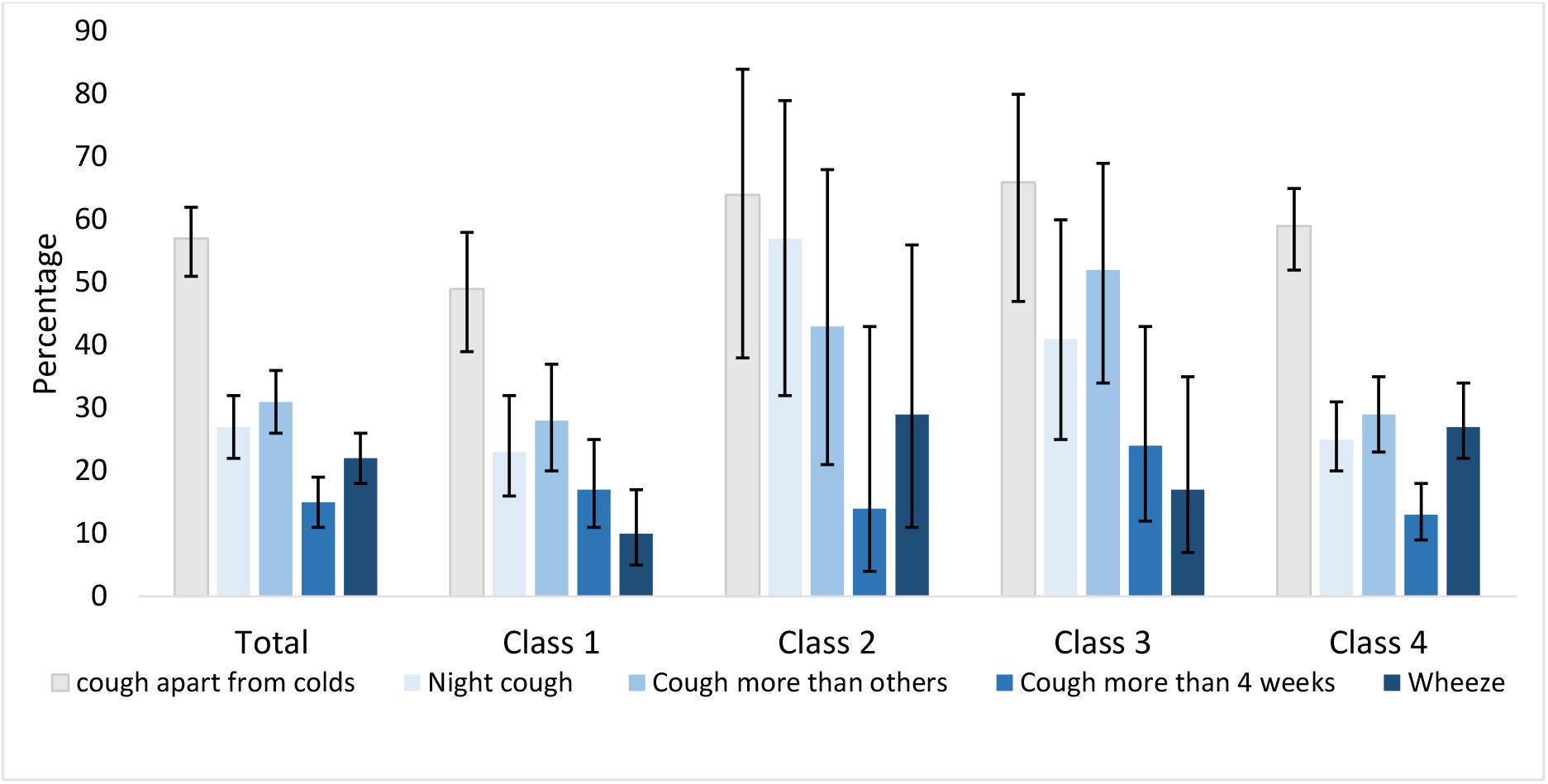
Prognostic outcomes after 1 year for the 4 cough phenotypes **Class 1**: Unspecific dry cough **Class 2**: Non-allergic infectious and night cough with snoring and otitis **Class 3:** Allergic dry night cough with snoring **Class 4:** Allergic cough

## 4. DISCUSSION

Our study applied LCA to identify phenotypes in children who cough without wheeze. We applied this method to a set of parent-reported cough-related symptoms and characteristics in a clinical-based study. We identified 4 cough phenotypes with the allergic cough phenotype as dominant. Upon validating the identified phenotypes, we found they differed with respect to parent-reported information about family histories and comorbidities, paediatric pulmonologist diagnoses, measurable traits, and respiratory prognosis 1 year later.

### 4.1 Strengths and limitations

Ours is the first study identifying cough phenotypes in a clinical cohort of children by applying a data-driven unsupervised learning approach to a wide range of cough symptoms and characteristics. We used broad inclusion criteria—answering positively to any question about cough—for our study population, which minimised the risk of missing some phenotypes. In a previous population-based study, we found only a partial overlap between positive answers to different cough questions, which likely reflects the existence of subgroups of children with differences in underlying pathophysiology.^[37]^ For phenotype definitions, we used only cough-related symptoms and characteristics easily collected in primary care settings to make our results applicable to all healthcare settings. Lastly, we used different ways to validate the 4 phenotypes, including parent-reported information, paediatric pulmonologist diagnoses, measurable traits, and 1-year prognosis to help ascertain clinical relevance. Our study also has some limitations. First, despite our use of a data-driven approach, selecting variables to include in the model always involves some degree of subjectivity. Second, our baseline questionnaire lacked information about signs and symptoms of some possible aetiologies of cough, such as gastroesophageal reflux and psychogenic and habit/tic cough, which thus could not serve as indicator variables to build the latent class model. Third, our study population is mainly representative of German- and French-speaking families in Switzerland and includes few recent immigrants. Finally, we did not externally validate our findings among an independent clinical population of children— an interest for future studies to determine if we identify similar phenotypes. However, we are unaware of a similar study for validation.

### 4.2 Comparison with other studies

Our study is the first of its type to use LCA for identifying cough phenotypes among a clinical population of children. We cannot fully compare our findings with population-based studies also using LCA or similar clustering techniques to distinguish cough phenotypes.^[15, 26^-^28, 38]^ We provide a summary of study designs and populations, inclusion criteria, indicator variables, identified phenotypes/trajectories, and validation processes from existing studies in Table S6. Spycher et al. and Weinmayr et al. included children who wheeze and used mostly wheeze-related symptoms with only few questions about cough as indicator variables in the LCA. ^[15, 27]^ Spycher et al. also included physiological measurements as indicator variables in the LCA.^[15]^ In contrast, we excluded children who wheezed, and as indicator variables, we used a rich set of only cough-related questions allowing us to identify more cough phenotypes and to describe them in more detail.

Divaret-Chauveau et al. and Rancière et al. investigated cough without cold and dry night cough trajectories, respectively.^[28, 38]^ Both studies used only cough questions collected at different time points to define cough trajectories/patterns, but they included children who wheeze; and the children were also younger (Table S6). In an analysis stratified by presence or absence of wheeze, Divaret-Chauveau et al. found physician-diagnosed asthma reported among a small proportion of children in all cough trajectories (2 to 12%) even in the absence of wheeze.^[28]^ Rancière et al. also reported physician-diagnosed asthma ever in all cough patterns (7 to 26%) but this included children who wheeze.^[38]^ In our study, parent-reported, physician-diagnosed asthma ever was also reported in all identified cough phenotypes, yet in much higher proportions (22 to 55%). This might be explained by the difference in study populations, since children visiting respiratory outpatient clinics likely present with more severe respiratory problems than children from the general population.

### 4.3 Interpretation and implication of findings

Our study distinguished 4 phenotypes of cough among children. Based only on parent-reported characteristics, distinctions between different phenotypes were not clear-cut, yet reflect the realities and difficulties of clinical practice when trying to establish underlying aetiologies of cough based on history alone with further examinations usually required. Nevertheless, interesting and clinically-relevant patterns, such as cough driven by allergies and upper airway pathology, correlating with several parent-reported characteristics, paediatric pulmonologist diagnoses, measurable traits and prognosis, emerged from the latent class model.

Interestingly, most children (70%) had phenotypes related to allergy (Phenotypes 3 and 4). In terms of clinical implications, children who cough with allergic symptoms possibly need allergy and asthma work-ups even in absence of reported wheeze. Divaret-Chauveau et al. also concluded as such; they found an association between different cough trajectories and asthma/allergic rhinitis.^[28]^ Similarly, Rancière et al. found an association between children with rising cough patterns—late and persistent coughers—and allergic comorbidity, including asthma, and atopy, independent of the presence of wheeze.^[38]^

Additionally, the relatively high proportion of children reporting wheeze at follow-up (overall: 22%, range 10 to 29% for all 4 phenotypes) supports the need for further asthma work-ups. Our statistical model identified specific groups of children who more often develop wheeze and possibly asthma in the future. Spycher et al. also found that risk of reporting wheeze at follow-up was higher among children of the persistent cough phenotype compared with the transient cough phenotype (25% vs 10%).^[15]^ Rancière et al. found prevalence of wheeze at follow-up lowest among preschool children with infrequent night cough (4%) compared with transient (8%) and rising pattern (consisting of late and persistent cough, 16%).^[38]^ Findings from these studies and ours highlight the importance of identifying subgroups of children with cough since their prognoses differ.

Another interesting finding of our study is the identification of 2 cough phenotypes potentially driven by upper airway problems—1 allergic (phenotype 3) and 1 non-allergic (phenotype 2). The presence of night cough and snoring, long duration of cough, and rhino- conjunctivitis in phenotype 3 suggests house dust mite allergy. Children with phenotype 2 also had a high probability of night-time cough and snoring, yet in contrast to children with phenotype 3, without an allergic component, with more frequent infections (colds, pneumonia, otitis), and rarely reported dry cough. Phenotype 2 possibly represents children with recurrent upper airway infections and adenoid hypertrophy or postnasal rhinorrhoea— a small group (4% of children) perhaps restricted by our study population of children aged 5 years and older when respiratory infections become less common.^[10]^ This group also had the lowest estimated median lung function parameters but the number of children in this group was small and estimates were subject to much uncertainty.

The second largest group (phenotype 1: unspecific dry cough) possibly represents children with the so-called *non-specific isolated cough*—a “dry cough in the absence of identifiable respiratory or known aetiology.”^[39]^ This group of children seems to have a more transient cough, as compared with the other phenotypes, their prognosis after 1 year was better with less cough and less wheeze.

## 5. CONCLUSION

The 4 cough phenotypes we identified by LCA revealed interesting clinical patterns, which may be helpful in clinical settings. Most children (70%) had a phenotype associated with allergies. Thus, most children who cough may benefit from allergy and asthma diagnostic work-ups even in absence of reported wheeze.

## Data Availability

The data supporting our study findings are available on reasonable request from the corresponding author.

## AUTHOR CONTRIBUTIONS

CK, ESLP, and MG designed and set-up the SPAC study. CK, MCM, ESLP, and MG conceptualised the analysis with input from SB, KH, AJ, PL, AM, NR, and BDS. SB, KH, AJ, PL, AM, NR, and CK recruited participants for the study. MCM, ESLP, and RM collected and entered data. MCM performed statistical analyses. BDS and ESLP supported statistical analyses. MCM drafted the manuscript with substantial input from CK. All authors interpreted the data, critically reviewed the manuscript, and approved the final version of the manuscript.

**ACKNOWLEDGEMENTS**

We thank the families who took part in the SPAC study, the research assistants (Natalie Messerli, Gia Thu Ly, Labinata Gjokaj and Meret Ryser) for helping with data collection and data entry, Kristin Marie Bivens, PhD (Institute of Social and Preventive Medicine, University of Bern) for her editorial contribution, PedNet Bern for supporting data collection in Bern, and the members of the SPAC study team.

SPAC study team members include D. Mueller-Suter, P. Eng, and A. Kuhn (Canton Hospital Aarau, Aarau, Switzerland); U. Frey, J. Hammer, A. Jochmann, D. Trachsel, and A. Oettlin (University Children’s Hospital Basel, Basel, Switzerland); P. Latzin, C. Abbas, M. Bullo, O. Fuchs, E. Kieninger, I. Korten, L. Krüger, B. Seyfried, F. Singer, and S. Yammine, C.de Jong (University Children’s Hospital Bern, Bern, Switzerland); C. Casaulta and P. Iseli (Children’s Hospital Chur, Chur, Switzerland); K. Hoyler (private pediatric pulmonologist, Horgen, Switzerland); S. Blanchon, S. Guerin, and I. Rochat (University Children’s Hospital Lausanne, Lausanne, Switzerland); N. Regamey, M. Lurà, M. Hitzler, A. Clavuot, K. Hrup, and J. Stritt (Canton Hospital Lucerne, Lucerne, Switzerland); J. Barben (Children’s Hospital St Gallen, St Gallen, Switzerland); O. Sutter (private pediatric practice, Worb, Bern, Switzerland); A. Moeller, A. Hector, K. Heschl, A. Jung, T. Schürmann, L. Thanikkel, and J. Usemann (University Children’s Hospital Zurich, Zurich, Switzerland); and C.E. Kuehni, C. Ardura- Garcia, D. Berger, T.Krasnova, R. Makhoul, M.C. Mallet, E. Pedersen, and M. Goutaki (University of Bern, Institute of Social and Preventive Medicine, Bern, Switzerland).

## REFERENCES

1. Chang AB, Robertson CF, Van Asperen PP, Glasgow NJ, Mellis CM, Masters IB, et al. A multicenter study on chronic cough in children : burden and etiologies based on a standardized management pathway. Chest. 2012;142(4):943–50.

2. Marchant JM, Newcombe PA, Juniper EF, Sheffield JK, Stathis SL, Chang AB. What is the burden of chronic cough for families? Chest. 2008;134(2):303–9.

3. Shields MD, Bush A, Everard ML, McKenzie S, Primhak R. BTS guidelines: Recommendations for the assessment and management of cough in children. Thorax. 2008;63 Suppl 3:iii1–iii15.

4. Cash H, Trosman S, Abelson T, Yellon R, Anne S. Chronic cough in children. JAMA Otolaryngol Head Neck Surg. 2015;141(5):417–23.

5. De Benedictis FM, Selvaggio D, De Benedictis D. Cough, wheezing and asthma in children: lesson from the past. Pediatric Allergy and Immunology. 2004;15(5):386–93.

6. Gao F, Gu Q-L, Jiang Z-D. Upper airway cough syndrome in 103 children. Chinese Medical Journal. 2019;132(6):653–8.

7. Asilsoy S, Bayram E, Agin H, Apa H, Can D, Gulle S, et al. Evaluation of chronic cough in children. Chest. 2008;134(6):1122–8.

8. Gedik AH, Cakir E, Torun E, Demir AD, Kucukkoc M, Erenberk U, et al. Evaluation of 563 children with chronic cough accompanied by a new clinical algorithm. Ital J Pediatr. 2015;41:73.

9. Yu X, Kong L, Jiang W, Dai Y, Wang Y, Huang L, et al. Etiologies associated with chronic cough and its clinical characteristics in school-age children. Journal of thoracic disease. 2019;11(7):3093–102.

10. Marchant JM, Masters IB, Taylor SM, Cox NC, Seymour GJ, Chang AB. Evaluation and outcome of young children with chronic cough. Chest. 2006;129(5):1132–41.

11. Chang AB, Glomb WB. Guidelines for evaluating chronic cough in pediatrics: ACCP evidence-based clinical practice guidelines. Chest. 2006;129(1 Suppl):260s–83s.

12. Chang AB, Oppenheimer JJ, Irwin RS. Managing Chronic Cough as a Symptom in Children and Management Algorithms: CHEST Guideline and Expert Panel Report. Chest. 2020;158(1):303–29.

13. Morice AH, Millqvist E, Bieksiene K, Birring SS, Dicpinigaitis P, Domingo Ribas C, et al. ERS guidelines on the diagnosis and treatment of chronic cough in adults and children. The European respiratory journal. 2020;55(1).

14. Mori M, Krumholz HM, Allore HG. Using Latent Class Analysis to Identify Hidden Clinical Phenotypes. JAMA. 2020;324(7):700–1.

15. Spycher BD, Silverman M, Brooke AM, Minder CE, Kuehni CE. Distinguishing phenotypes of childhood wheeze and cough using latent class analysis. Eur Respir J. 2008;31(5):974–81.

16. Ross MK, Eckel SP, Bui AAT, Gilliland FD. Asthma clustering methods: a literature-informed application to the children’s health study data. J Asthma. 2022;59(7):1305–18.

17. Spycher BD, Silverman M, Kuehni CE. Phenotypes of childhood asthma: are they real? Clin Exp Allergy. 2010;40(8):1130–41.

18. Lee E, Lee SH, Kwon JW, Kim YH, Yoon J, Cho HJ, et al. Persistent asthma phenotype related with late-onset, high atopy, and low socioeconomic status in school-aged Korean children. BMC Pulm Med. 2017;17(1):45.

19. Brew BK, Chiesa F, Lundholm C, Örtqvist A, Almqvist C. A modern approach to identifying and characterizing child asthma and wheeze phenotypes based on clinical data. PLoS One. 2019;14(12):e0227091.

20. Oksel C, Granell R, Haider S, Fontanella S, Simpson A, Turner S, et al. Distinguishing Wheezing Phenotypes from Infancy to Adolescence. A Pooled Analysis of Five Birth Cohorts. Ann Am Thorac Soc. 2019;16(7):868–76.

21. Fitzpatrick AM, Bacharier LB, Guilbert TW, Jackson DJ, Szefler SJ, Beigelman A, et al. Phenotypes of Recurrent Wheezing in Preschool Children: Identification by Latent Class Analysis and Utility in Prediction of Future Exacerbation. J Allergy Clin Immunol Pract. 2019;7(3):915–24.e7.

22. Fitzpatrick AM, Bacharier LB, Jackson DJ, Szefler SJ, Beigelman A, Cabana M, et al. Heterogeneity of Mild to Moderate Persistent Asthma in Children: Confirmation by Latent Class Analysis and Association with 1-Year Outcomes. J Allergy Clin Immunol Pract. 2020;8(8):2617–27.e4.

23. Ferrante G, Fondacaro C, Cilluffo G, Dones P, Cardella F, Corsello G. Identification of bronchiolitis profiles in Italian children through the application of latent class analysis. Ital J Pediatr. 2020;46(1):147.

24. Niu H, Chang AB, Oguoma VM, Wang Z, McCallum GB. Latent class analysis to identify clinical profiles among indigenous infants with bronchiolitis. Pediatr Pulmonol. 2020;55(11):3096–103.

25. Petrarca L, Nenna R, Di Mattia G, Frassanito A, Castro-Rodriguez JA, Rodriguez Martinez CE, et al. Bronchiolitis phenotypes identified by latent class analysis may influence the occurrence of respiratory sequelae. Pediatr Pulmonol. 2022;57(3):616–22.

26. Spycher BD, Silverman M, Pescatore AM, Beardsmore CS, Kuehni CE. Comparison of phenotypes of childhood wheeze and cough in 2 independent cohorts. J Allergy Clin Immunol. 2013;132(5):1058–67.

27. Weinmayr G, Keller F, Kleiner A, du Prel JB, Garcia-Marcos L, Batllés-Garrido J, et al. Asthma phenotypes identified by latent class analysis in the ISAAC phase II Spain study. Clin Exp Allergy. 2013;43(2):223–32.

28. Divaret-Chauveau A, Mauny F, Hose A, Depner M, Dalphin ML, Kaulek V, et al. Trajectories of cough without a cold in early childhood and associations with atopic diseases. Clin Exp Allergy. 2022.

29. Pedersen ESL, de Jong CCM, Ardura-Garcia C, Barben J, Casaulta C, Frey U, et al. The Swiss Paediatric Airway Cohort (SPAC). ERJ Open Res. 2018;4(4).

30. Ferris BG. Epidemiology Standardization Project (American Thoracic Society). Am Rev Respir Dis. 1978;118(6 Pt 2):1–120.

31. Asher MI, Keil U, Anderson HR, Beasley R, Crane J, Martinez F, et al. International Study of Asthma and Allergies in Childhood (ISAAC): rationale and methods. Eur Respir J. 1995;8(3):483–91.

32. Clifford RD, Radford M, Howell JB, Holgate ST. Prevalence of respiratory symptoms among 7 and 11 year old schoolchildren and association with asthma. Arch Dis Child. 1989;64(8):1118–25.

33. de Onis M, Onyango AW, Borghi E, Siyam A, Nishida C, Siekmann J. Development of a WHO growth reference for school-aged children and adolescents. Bull World Health Organ. 2007;85(9):660–7.

34. Quanjer PH, Stanojevic S, Cole TJ, Baur X, Hall GL, Culver BH, et al. Multi-ethnic reference values for spirometry for the 3-95-yr age range: the global lung function 2012 equations. Eur Respir J. 2012;40(6):1324–43.

35. Araya-Cloutier CV J; Van de Schans, M; Hageman, J; Schaftenaar, G. ATS/ERS recommendations for standardized procedures for the online and offline measurement of exhaled lower respiratory nitric oxide and nasal nitric oxide, 2005. Am J Respir Crit Care Med. 2005;171(8):912–30.

36. Weller BEB N.K; Faubert, S.J. Latent Class Analysis: A Guide to Best Practice. Journal of Black Pyschology. 2020;46(4):287–311.

37. Mallet MCM R; Pedersen, E.S.L; Ardura-Garcia, C; Gaillard, E.A; Latzin, P; Moeller, A; Kuehni, C.E. Prevalence of childhood cough in epidemiological studies depends on the question used: findings from two population-based studies. Swiss Med Wkly. 2023; 153:40044.

38. Rancière F, Nikasinovic L, Momas I. Dry night cough as a marker of allergy in preschool children: the PARIS birth cohort. Pediatr Allergy Immunol. 2013;24(2):131–7.

39. Chang AB. Chronic non-specific cough in children. Paediatrics and Child Health. 2008;18(7):333–9.

