## Supplementary material for "Phenotypes of cough in children: a latent class analysis": Online supplements Appendix S1

### Supplementary methods

#### 2.4 Definition of variables to characterise and validate identified phenotypes

***Parent-reported characteristics***

We assessed how the identified phenotypes differed with respect to the following parent-reported information from the baseline questionnaire:

- *sociodemographic characteristics* (sex, age, country of origin, and parental education)
- *environmental exposure* (parental smoking)
- *family history* (asthma or inhaler use, hay fever, and chronic cough)
- *comorbidities in the child's lifetime* (previous wheeze, physician-diagnosed asthma, hay fever, and eczema).

***Paediatric pulmonologist diagnoses***

We compared phenotypes with diagnoses given by paediatric pulmonologists who were unaware of the latent class analysis results and grouped them into diagnostic categories. A child could have several diagnoses, such as

- *asthma and asthma-like diagnosis* (chronic asthma, episodic asthma, exercise-induced asthma, wheeze, recurrent bronchitis, bronchial hyperreactivity, exercise-induced wheeze)
- *chronic cough* (recurrent cough of unknown aetiology, increased vulnerability to viral infections, post-infectious cough, psychogenic cough, chronic obstructive rhinopathy

with posterior rhinorrhea, cough hypersensitivity syndrome, protracted bacterial bronchitis, gastroesophageal reflux disease)

- *exercise-related problems* (dysfunctional breathing, hyperventilation syndrome, stridor, sighing, vocal cord dysfunction, induced laryngeal obstruction, deconditioning)
- *chronic snoring* (adenoid hypertrophy, tonsil hypertrophy, obstructive sleep apnea and unspecific snoring)
- *rhinitis or rhino-conjunctivitis* and *allergic sensitisation/atopy*

### Supplementary Tables and Figures

**Table S1:** Definitions, answer categories, and percentages of missing of the indicator variables used to build the latent class analysis model to identify cough phenotypes among children (study population, n=531)

| Variable | Question | Answer categories , N(%) | Dichotomisation, N(%) | Missing |
|---|---|---|---|---|
| Number of colds | How many colds did your child have in the past 12 months?[1] | (i) None, 9 (2)<br>(ii) 1-3, 298 (56)<br>(iii) 4-6, 146 (28)<br>(iv) 7-9, 50 (9)<br>(v) ≥ 10, 11 (2)<br>(vi) Missing, 17 (3) | (i) 0-6, 470 (89)<br>(ii) ≥ 7, 61 (11) | Missing recoded using the median number of colds (1–3) |
| Pneumonia | Did your child have a pneumonia in the last 12 months?[2] | (i)None, 495 (93)<br>(ii) Yes, once, 27 (5)<br>(iii) Yes, several times, 4 (1)<br>(iv) Missing, 5 (1) | (i)No, 500 (94)<br>(ii)Yes, 31 (6) | Missing recoded as “no” |
| Otitis media | Did your child have a middle ear infection in the past 12 months?[2] | (i)None, 472 (89)<br>(ii) Yes, once, 33 (6)<br>(iii) Yes, several times, 14 (3)<br>(iv) Missing, 12 (2) | (i)No, 484 (91)<br>(ii)Yes, 47 (9) | Missing recoded as “no” |
| Snoring | During the past 12 months, has your child sometimes snored?[1] | (i)No, 174 (33)<br>(Ii)Only with colds, 159 (30)<br>(Iii)Also without colds, 144 (27)<br>(iv) Almost always, 42 (8)<br>(v) Missing, 12 (2) | (i)No/only with colds, 345 (65)<br>(ii)Without colds/almost always, 186 (35) | Missing recoded as “no” |
| Rhino conjunctivitis | In the past 12 months, has your child had a problem with sneezing, or a runny, or blocked nose when he/she did not have a cold or the flu? If yes, in the past 12 months, has this nose problem been accompanied by itchy-watery eyes? [3] | (i)No, 301 (57)<br>(ii)Yes, 195 (37)<br>(iii) Missing, 35 (7) | (i)No, 336 (63)<br>(ii)Yes, 195 (37) | Missing recoded as “no” |
| Allergic cough triggers | Which of the following situation has caused your child to experience coughing or wheezing in the last 12 months?[1] | | | |
| | Pollen | (i)Never, 204 (38)<br>(ii)Sometimes, 149 (28)<br>(iii)Often, 101 (19)<br>(iv)Missing, 77 (15) | (i)No, 281 (53)<br>(ii)Yes, 250 (47) | Missing recoded as “no” |
| | Animal | (i)Never, 327 (62)<br>(ii)Sometimes, 79 (15)<br>(iii)Often, 36 (7)<br>(iv)Missing, 89 (17) | (i)No, 416 (78)<br>(ii)Yes, 115 (22) | Missing recoded as “no” |
| | House dust | (i)Never, 265 (50)<br>(ii)Sometimes, 118 (22)<br>(iii)Often, 49 (9)<br>(iv)Missing, 99 (19) | (i)No, 364 (69)<br>(ii)Yes, 167 (31) | Missing recoded as “no” |
| | Number of allergic cough triggers | (i)0, 213(49)<br>(ii)1, 158 (30)<br>(iii)2, 106 (20)<br>(iv)3, 54 (10) | Use as ‘count’ variable | |
| Night cough | In the last 12 months has your child had a dry cough at | (i)No, 246 (46)<br>(ii)Yes, 276 (52) | (i)No, 255(48)<br>(ii)Yes, 276 (52) | Missing recoded as “no” |

| | | | | |
|---|---|---|---|---|
| | night, apart from a cough associated with a cold or a chest infection [3] | (iii) Missing, 9 (2) | | |
| Type of cough | Most of the time, is your child's cough rather dry or wet?[1] | (i)Mostly dry, 253 (48)<br>(ii)Mostly wet, 53 (10)<br>(iii)Both, 221 (42)<br>(vi)Missing, 4 (1) | (i)Mostly dry → 253 (48)<br>(ii)Most wet/both → 274(52)<br>Missing: 4 (1) | |
| Cough > 2 months | Has your child had a cough in the past 12 months that lasted more than 2 months in a row?[1] | (i)No, 434 (82)<br>(ii)Yes, 79 (15)<br>(iii)Missing, 18 (3) | (i)No → 452 (85)<br>(ii)Yes → 79 (15) | Missing recoded as "no" |

**Table S2:** Frequency of diagnostic tests done and follow-up questionnaires completed, in total, and based on the 4 classes

| **Class** | | **1** | **2** | **3** | **4** | |
|---|---|---|---|---|---|---|
| **Class description** | All | Unspecific dry cough | Non-allergic infectious and night cough with snoring and otitis | Allergic dry night cough with snoring | Allergic cough | |
| | (N=531) | (N=145) | (N=18) | (N=45) | (N=323) | |
| | N (%) | N (%) | N (%) | N (%) | N (%) | p-value |
| Spirometry parameters | | | | | | |
| FEV1 | 451 (85) | 125 (86) | 16 (89) | 41 (91) | 269 (83) | 0.487 |
| FVC | 440 (83) | 125 (86) | 16 (89) | 38 (84) | 261 (81) | 0.450 |
| FEV1/FVC | 436 (82) | 122 (84) | 16 (89) | 38 (84) | 260 (81) | 0.635 |
| FEF 25–75 | 316 (60) | 94 (65) | 11 (61) | 25 (56) | 186 (58) | 0.474 |
| Bronchodilator response test | 239 (45) | 73 (50) | 10 (56) | 14 (31) | 142 (44) | 0.105 |
| FeNO | | | | | | |
| Online and offline | 479 (90) | 126 (87) | 15 (83) | 42 (93) | 296 (92) | 0.220* |
| Online only | 450 (85) | 116 (80) | 13 (72) | 37 (82) | 284 (88) | 0.059 |
| Allergy tests | 353 (66) | 93 (64) | 13 (72) | 32 (71) | 215 (67) | 0.787 |
| Completed first year follow-up questionnaire | 360 (68) | 101 (70) | 14 (78) | 29 (64) | 216 (67) | 0.704 |

FEV1: forced expiratory volume in 1 second; FVC: forced vital capacity, FEF 25–75: forced expiratory flow at 25–75% of the vital capacity; FeNO: fractional exhaled nitric oxide; *p*-values calculated using Chi-square tests, **p*-value calculated using Fisher's exact test

**Figure S1:** Flowchart of included participants

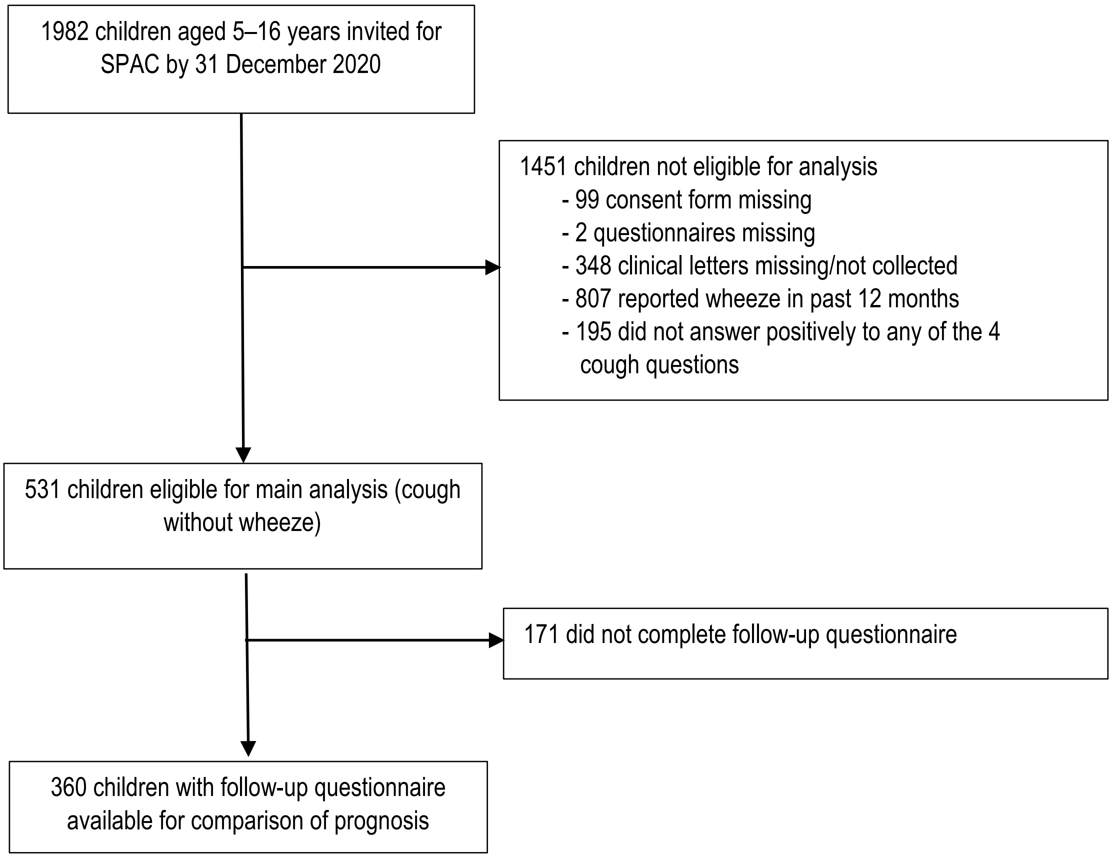

**Table S3:** Statistical fit indices of the different latent class models of cough phenotypes (N=531)

| Models | Log-likelihood | AIC | BIC |
|---|---|---|---|
| 2 classes | -2780.6 | 5599.2 | **5680.421** |
| 3 classes | -2761.263 | 5580.526 | 5704.494 |
| 4 classes | -2741.936 | **5561.873** | 5728.589 |
| 5 classes | -2732.687 | 5563.373 | 5772.836 |
| 6 classes | -2725.704 | 5569.408 | 5821.619 |
| 7 classes | -2719.277 | 5566.555 | 5840.139 |
| 8 classes | -2717.439 | 5592.878 | 5930.584 |
| 9 classes | -2707.123 | 5590.247 | 5966.426 |
| 10 classes | -2701.068 | 5598.136 | 6017.062 |

AIC: Akaike information criterion; BIC: Bayesian information criterion; the lowest values of AIC and BIC represents the best fitting models; Values in **bold** are the lowest values for AIC and BIC

**Table S4:** Estimated class sizes and estimated probabilities of cough-related symptoms and characteristics of the 4-class models identified by latent class analysis based on the lowest Akaike Information Criterion (N=531)

| | Class 1 | Class 2 | Class 3 | Class 4 |
|---|---|---|---|---|
| ***Latent class probabilities*** | 0.25 | 0.04 | 0.09 | 0.61 |
| ***Item response probabilities*** | | | | |
| Frequent colds ≥7/year | 0.15 | 0.29 | 0.18 | 0.08 |
| Pneumonia | 0.06 | 0.21 | 0.19 | 0.03 |
| Otitis media | 0.02 | **0.79** | 0.00 | 0.09 |
| Snoring (also without cold and almost always) | 0.18 | **0.84** | **0.68** | 0.34 |
| Rhino-conjunctivitis | 0.01 | 0.00 | **0.39** | **0.54** |
| Number of allergic triggers# | 0.18 | 0.38 | **1.34** | **1.33** |
| Night cough | 0.33 | **0.91** | **1.00** | 0.50 |
| Mostly dry cough | **0.51** | 0.26 | **0.71** | 0.45 |
| Cough > 2 months | 0.13 | 0.18 | **0.72** | 0.07 |

Width of the column represents relative sizes of classes. All the symptoms are parent-reported and are from the last 12 months; # represents estimated mean number of triggers (range 0–3) and calculated using Poisson. In **bold** are features that differ between classes.

**Legend: range of estimated class size and item response probabilities**

| | | | | |
|---|---|---|---|---|
| <0.20 | ≥0.20 to <0.40 | ≥0.40 to <0.60 | ≥0.60 to <0.80 | ≥0.80 |

**Legend: range of estimated mean number of allergic triggers**

| | | | | |
|---|---|---|---|---|
| 0 | >0 to <1 | ≥1 to <2 | ≥2 to <3 | 3 |

**Class 1**: Unspecific dry cough
**Class 2**: Non-allergic infectious and night cough with snoring and otitis
**Class 3:** Allergic dry night cough with snoring
**Class 4**: Allergic cough

**Table S5:** Estimated class sizes and estimated probabilities of cough-related symptoms and characteristics and estimated class sizes of the two-class model identified by latent class analysis based on the lowest Bayesian Information Criterion (N=531)

| | Latent class | |
|---|---|---|
| | **Class 1** | **Class 2** |
| ***Latent class probabilities*** | 0.73 | 0.27 |
| ***Item response probabilities*** | | |
| Frequent colds ≥7/year | 0.09 | 0.17 |
| Pneumonia | 0.05 | 0.08 |
| Otitis media | 0.09 | 0.07 |
| Snoring (also without cold and almost always) | 0.39 | 0.24 |
| Rhinoconjunctivitis | 0.51 | 0.01 |
| Number of allergic triggers# (mean count) | 1.32 | 0.17 |
| Night cough | 0.57 | 0.39 |
| Mostly dry cough | 0.48 | 0.49 |
| Cough > 2 months | 0.15 | 0.15 |

All the symptoms are parent-reported and are from the last 12 months; # represents mean number of triggers (range 0–3) and calculated using Poisson

**Figure S2:** Distribution of spirometry parameters z-scores (FEV1, FVC, FEV1/FVC and FEF 25-75) based on the 4 identified cough phenotypes

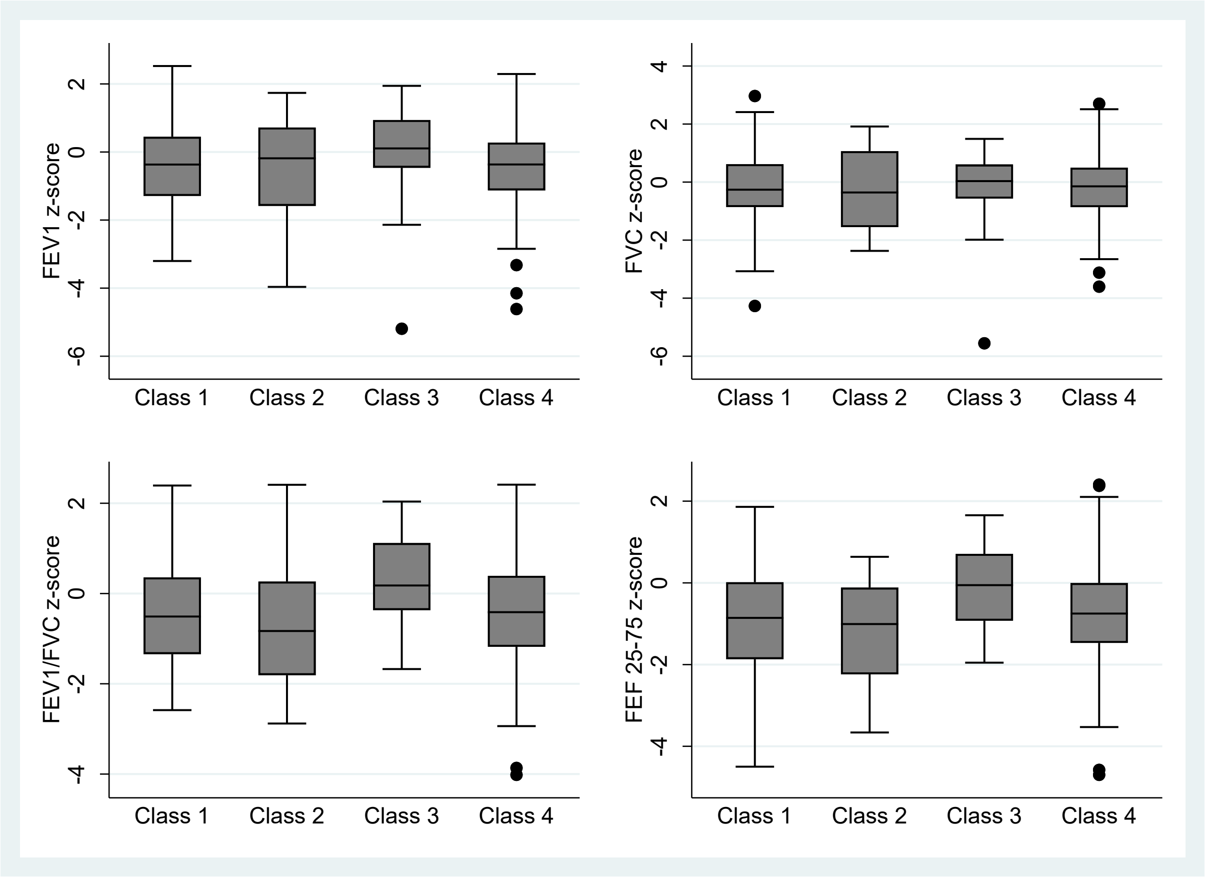


FEV1: forced expiratory volume in 1 second; FVC: forced vital capacity, FEF 25–75: forced expiratory flow at 25–75% of the vital capacity

**Class 1**: Unspecific dry cough
**Class 2**: Non-allergic infectious and night cough associated with snoring and otitis
**Class 3:** Allergic dry night cough associated with snoring and allergies
**Class 4**: Allergic cough

**Table S6:** Summary of studies looking at cough phenotypes or trajectories using a clustering approach

| Study and year of publication | Study design and population | Inclusion criteria | Clustering method used and indicator variables | Phenotypes/ trajectories identified | Variable used for characterisation/ validation of phenotypes | External validation |
|---|---|---|---|---|---|---|
| Spycher et al., 2008 [4] | Population-based longitudinal study in UK<br><br>Preschool children<br><br>First survey in 1990 and second survey in 1992–1994 | Positive answer to attacks of wheezing ever and/or usually have a cough apart from colds in one or both surveys.<br><br>A control group (i.e. asymptomatic children in both surveys) was included. | Clustering method: Latent class analysis (LCA)<br><br>Indicator variables:<br>-Wheeze ever<br>-Attacks in last 12 months<br>-Attacks with shortness of breath<br>-Triggers of wheeze<br>-Season with most frequent attacks<br>-Time of day with worse attacks<br>-Wakened by cough at night<br>-Cough only with colds/apart from colds<br>-Sex<br>-Age<br>-Skin prick tests<br>-FEV0.5<br>-Bronchial responsiveness | 5 phenotypes:<br>(i)Persistent cough<br>(ii)Transient cough<br>(iii)Atopic persistent wheeze<br>(iv)Nonatopic persistent wheeze<br>(v)Transient viral wheeze | Prognosis after 5 and 10 years (wheeze, frequent wheeze, bronchodilator use and cough without colds) | Validation in an external independent population-based cohort [5] |
| Weinmayr et al., 2013[6] | Cross-sectional international study on allergies and asthma in childhood (ISAAC)-Phase 2<br><br>School children aged 8–12 years from Spain | Children with a complete set of parental responses on respiratory symptoms. | Clustering method: LCA<br><br>Indicator variables:<br>-Wheeze past year<br>-Wheeze if exercise<br>-Wheeze if no exercise<br>-Wheeze with a cold<br>-Wheeze if no cold<br>-Dry cough at night<br>-Coughed up phlegm with colds<br>-Coughed up phlegm without colds<br>-Coughed up phlegm frequently<br>-Ever woken with shortness of breath<br>-Ever woken with tightness of the chest | 6 phenotypes:<br>(i)No respiratory symptoms<br>(ii)Cough during colds<br>(iii)Chronic cough and phlegm<br>(iv)Nocturnal breathlessness<br>(v)Wheeze only with colds<br>(vi)Wheeze without colds, with cough<br>(vii)Wheeze without colds, without cough | -Allergic sensitization (skin prick tests and IgE)<br>-Lung function (spirometry)<br>-Bronchial hyperresponsiveness<br>-Demographic characteristics<br>-Clinical symptoms of rhinitis<br>-Examined eczema<br>-Parental history of asthma<br>-Disease severity | No |

| | | | | | | |
|---|---|---|---|---|---|---|
| Divaret-Chauveau et al., 2022 [7] | European prospective birth cohort (pregnant women recruited in third trimester of pregnancy)<br><br>Parental questionnaires completed at 1, 1.5, 2, 3 4, 5, 6 and 10 years | Children enrolled in the birth cohort and with at least 6 visits | Clustering method: LCA<br><br>Indicator variables (occurrence at 1, 1.5, 2, 3, 4, 5, 6 and 10 years old):<br><br>-Cough without a cold<br>-Cough at night without a cold<br>-Cough attack without a cold caused by one of the following factors: physical exercise, excitation, change of temperature | 9 trajectories of cough without cold<br>(i)Never/infrequent cough<br>(ii) 5 acute transient cough<br>(iii)Moderate transient cough<br>(iv)Late persistent cough<br>(v)Early persistent cough | -Characteristics of study population (growing up on a farm, parental history of atopy, gender)<br>-Atopic diseases and sensitization (unremitting wheeze, physician-diagnosed asthma, allergic rhinitis, atopic dermatitis, food allergy, sensitization to perennial, seasonal or food allergens) | No |
| Rancière et al., 2013 [8] | Birth cohort of full-term, healthy infants from 5 maternity hospitals in PARIS<br><br>Children from birth to age 4 years | All children with a complete report of dry night cough at 1, 2, 3 and 4 years) | Clustering method: k-means clustering<br><br>Indicator variables (occurrence at 1, 2, 3 and 4 years)<br>-Dry night cough | 3 trajectories of dry night cough<br>(i)Never/infrequent pattern<br>(ii)Transient pattern<br>(iii)Rising pattern (including persistent and late coughers) | -Blood markers of atopy (total IgE, eosinophilia, sensitisation to allergens)<br>-reported allergic symptoms (wheezing, rhinoconjunctivitis like symptoms, atopic dermatitis-like symptoms)<br>-reported physician-diagnosed diseases (asthma, eczema, hay fever, or food allergy) | No |

### References


1. Mozun R, Kuehni CE, Pedersen ESL, Goutaki M, Kurz JM, de Hoogh K, et al. LuftiBus in the school (LUIS): a population-based study on respiratory health in schoolchildren. Swiss Med Wkly. 2021;151:w20544.
2. Kuehni CE, Brooke AM, Strippoli MP, Spycher BD, Davis A, Silverman M. Cohort profile: the Leicester respiratory cohorts. Int J Epidemiol. 2007;36(5):977-85.
3. Asher MI, Keil U, Anderson HR, Beasley R, Crane J, Martinez F, et al. International Study of Asthma and Allergies in Childhood (ISAAC): rationale and methods. Eur Respir J. 1995;8(3):483-91.
4. Spycher BD, Silverman M, Brooke AM, Minder CE, Kuehni CE. Distinguishing phenotypes of childhood wheeze and cough using latent class analysis. Eur Respir J. 2008;31(5):974-81.
5. Spycher BD, Silverman M, Pescatore AM, Beardsmore CS, Kuehni CE. Comparison of phenotypes of childhood wheeze and cough in 2 independent cohorts. J Allergy Clin Immunol. 2013;132(5):1058-67.
6. Weinmayr G, Keller F, Kleiner A, du Prel JB, Garcia-Marcos L, Batllés-Garrido J, et al. Asthma phenotypes identified by latent class analysis in the ISAAC phase II Spain study. Clin Exp Allergy. 2013;43(2):223-32.
7. Divaret-Chauveau A, Mauny F, Hose A, Depner M, Dalphin ML, Kaulek V, et al. Trajectories of cough without a cold in early childhood and associations with atopic diseases. Clin Exp Allergy. 2022.
8. Rancière F, Nikasinovic L, Momas I. Dry night cough as a marker of allergy in preschool children: the PARIS birth cohort. Pediatr Allergy Immunol. 2013;24(2):131-7.
